# An Upsurge of SARS CoV-2 B.1.1.7 Variant in Pakistan

**DOI:** 10.1101/2021.02.26.21252562

**Authors:** Massab Umair, Muhammad Salman, Zaira Rehman, Nazish Badar, Abdul Ahad, Aamer Ikram

## Abstract

The emergence of a more transmissible variant of SARS-CoV-2 (B1.1.7) in the United Kingdom (UK) during late 2020 has raised major public health concerns. Several mutations have been reported in the genome of the B.1.1.7 variant including the N501Y and 69-70deletion in the Spike that has implications on virus transmissibility and diagnostics. Although the B.1.1.7 variant has been reported from several countries, only two cases have been identified through whole-genome sequencing from Pakistan. We used a two-step strategy for the detection of B.1.1.7 with initial screening through ThermoFisher TaqPath™ SARS-CoV-2 kit followed by partial sequencing of Spike (S) gene of samples having spike gene target failure (SGTF) on real-time PCR. From January 01, 2021, to February 21, 2021, a total of 2,650 samples were tested for the presence of SARS-CoV-2 using TaqPath™ kit and 70.4% (n=1,867) showed amplification of all the 3 genes (S, N, and ORF). Notably, 29.6% (n=783) samples had the spike gene target failure (SGTF). The SGTF cases were detected at a low frequency during the first three weeks of January (n=10, n=13, and n=1 respectively) however, the cases started to increase in the last week. During February, 726 (93%) cases of SGTF was reported with a peak (n=345) found during the 3rd week. Based on the partial sequencing of spike gene of SGTF samples (n=15), 93% (n=14) showed the characteristic N501Y, A570D, P681H, and T716I mutations found in the B.1.1.7 variant. Our findings highlight the high prevalence of B.1.1.7 in Pakistan and warrant large scale genomic surveillance and strengthening of laboratory network in the country.

## Introduction

The last few months of 2020 have witnessed the emergence of different variants of severe acute respiratory syndrome coronavirus 2 (SARS CoV-2) having characteristic mutations in their genome [1]. One such variant is the B.1.1.7 (VOC 202012/01) which was first detected in the United Kingdom (UK) during November 2020, later rapidly became one of the most prevalent form of SARS-CoV-2 in the country. Due to its high transmissibility rate (about 50-70%) [2], the variant has now spread to 93 countries as of February 21, 2021 [3].

The B.1.1.7 harbors 17 amino acid changes mostly in the Spike (S) protein (69-70del, 144del, N501Y, A570D, P681H, T716I, S982A and D1118H) [4]. Mutations in the S protein of SARS-CoV-2, which is involved in viral entry into host cells can potentially impact virus transmissibility, diagnostic capability and pathogenesis. Of particular note is the N501Y change in the receptor binding domain (RBD) that is directly involved in interactions with the human angiotensin-converting enzyme 2 (hACE2) receptor [5]. The N501Y mutation can lead to enhanced viral binding efficiency with hACE2 thus rendering the virus more transmissible [6, 7]. Similarly, the 69-70 deletion (del69-70) in the S gene may increase cell infectivity and lead to diagnostic failure in real-time PCR amplification kits targeting this region [8, 9]. One such assay is the ThermoFisher TaqPath™ SARS-CoV-2 kit that targets three genes of the virus including the S gene [10]. With this method, the del69-70 in the S gene of SARS-CoV-2 results in the spike gene target failure (SGTF) on real-time PCR that has been used as a proxy for the detection of B.1.1.7 variant [9, 11]. Recently, the European CDC and US FDA also recommended that SGFT can be used as signal for early identification of del69-70 that can be further investigated to track mutant virus [12].

Currently, the genomic surveillance of SARS-CoV-2 in Pakistan is limited to few laboratories having specialized facilities and expertise that has resulted in uncertainty about the introduction, geographic spread and community transmission of variants such as the B.1.1.7. As of 26 February 2021, only two cases of B.1.1.7 have been reported from Pakistan through whole-genome sequencing by the National Institute of Health [13]. We aimed to investigate the prevalence of B.1.1.7 in Pakistan using the SGTF method followed by confirmation of a subset of samples through partial sequencing of S gene (encompassing the RBD).

## Material & Methods

Oropharyngeal samples collected from patients suspected of COVID-19 received at the Department of Virology, National Institute of Health were included in the study. RNA was extracted from the samples using MagMAX(tm) Viral/Pathogen Nucleic Acid Isolation Kit and KingFisher ™ Flex Purification System (ThermoFisher). For the detection of SARS-CoV-2, TaqPath ™ COVID-19 CE-IVD RT-PCR Kit (ThermoFisher) that targets three genes (ORF1ab, N and S) was used. Samples with Spike Gene Target Failure (SGTF) were selected for the partial sequencing of the spike gene (S). The RT-PCR was carried out using the Qiagen™ OneStep RT-PCR kit according to the manufacturer’s instructions. The primers used for the partial sequencing of S gene had been reported previously [14] and the PCR was optimized with Qiagen™ OneStep RT-PCR kit. Following thermal profile was used for the amplification of spike gene fragment: 50°C for 30 min, 95°C for 15 min followed by 40 cycles of 94°C for 1 min, 55°C for 30 sec and 72°C for 1 min. PCR was completed with a final extension at 72°C for 10 min. The amplified product was sequenced using the BigDye™ Terminator v3.1 Cycle Sequencing kit on ABI3500xL genetic analyzer (ThermoFisher). All the sequences are submitted to the GISAID under the accession numbers: (EPI_ISL_1072998, EPI_ISL_1073001, EPI_ISL_1073004, EPI_ISL_1073012, EPI_ISL_1073015, EPI_ISL_1073021, EPI_ISL_1073023 - EPI_ISL_1073028, EPI_ISL_1073030 - EPI_ISL_1073033).

## Results and Discussion

During January 01, 2021 to February 21, 2021, a total of 2,650 samples were tested for the presence of SARS-CoV-2 on real-time PCR using TaqPath™ kit. Among the total samples, 70.4% (n=1,867) showed amplification of all the 3 genes (S, N, and ORF). Notably, 783 (29.6%) samples had the spike gene target failure (SGTF). These SGTF cases were detected at a low frequency during the first three week of January (n=10, n=13 and n=1 respectively) however, the cases started to increase in the last week. During February, 726 (93%) cases of SGTF were reported with a peak (n=345) found during the 3^rd^ week (Fig. 1).

**Figure 1:**
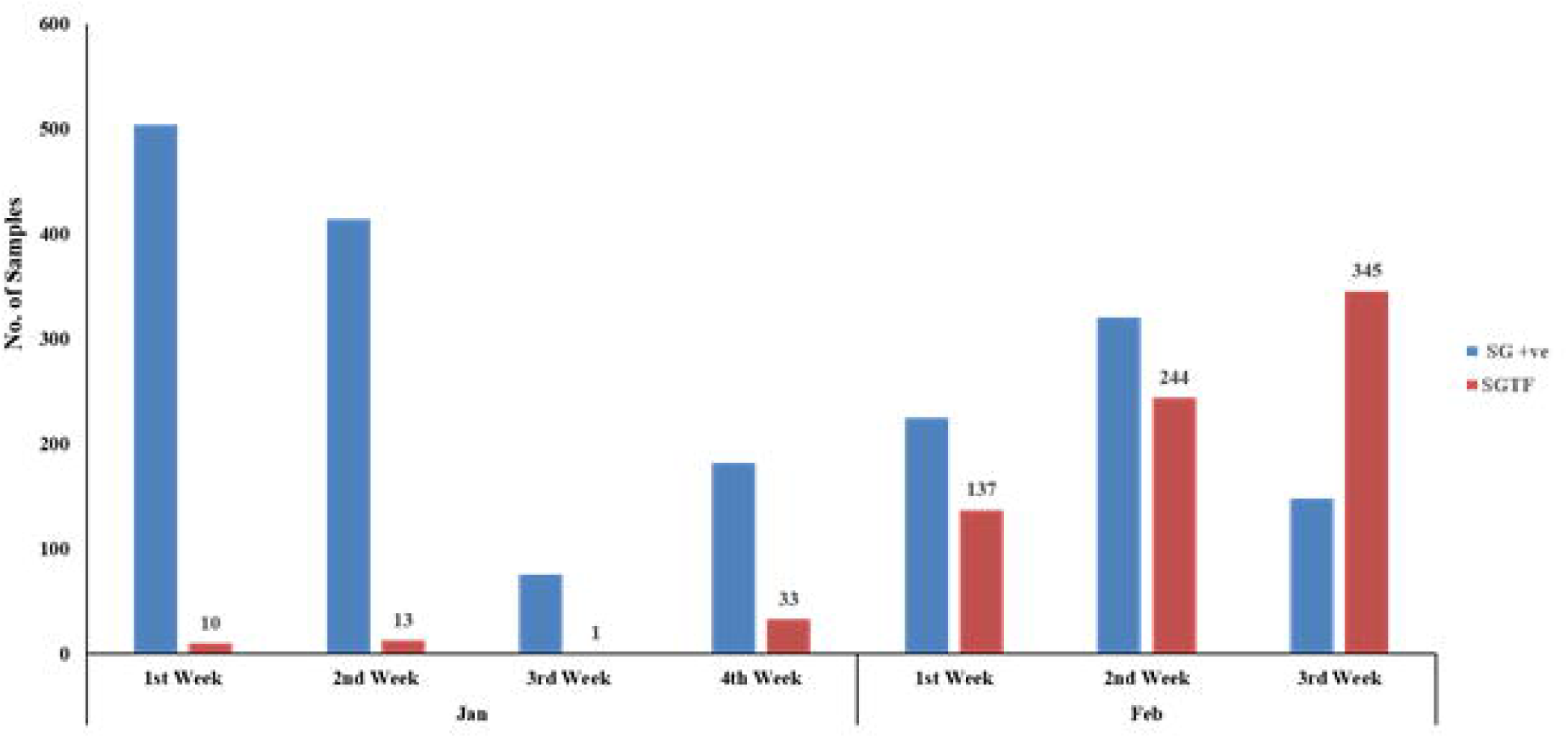
The distribution of COVID-19 patients according to the Δ69-70 deletion during January and first three weeks of February, 2021. TaqPath™ RT-PCR kit was used for detection of SGTF in SARS_CoV-2 patients. SG +ve: spike detection, SGTF: spike not detected.

Based on the partial sequencing of spike gene of SGTF samples (n=15), 14 showed the characteristic N501Y, A570D, P681H, and T716I mutations found in the B.1.1.7 strains. However, one of the SGTF sample (4490AB) does not harbor any characteristics B1.1.7 mutation in spike gene (Table 1).

**Table 1:**
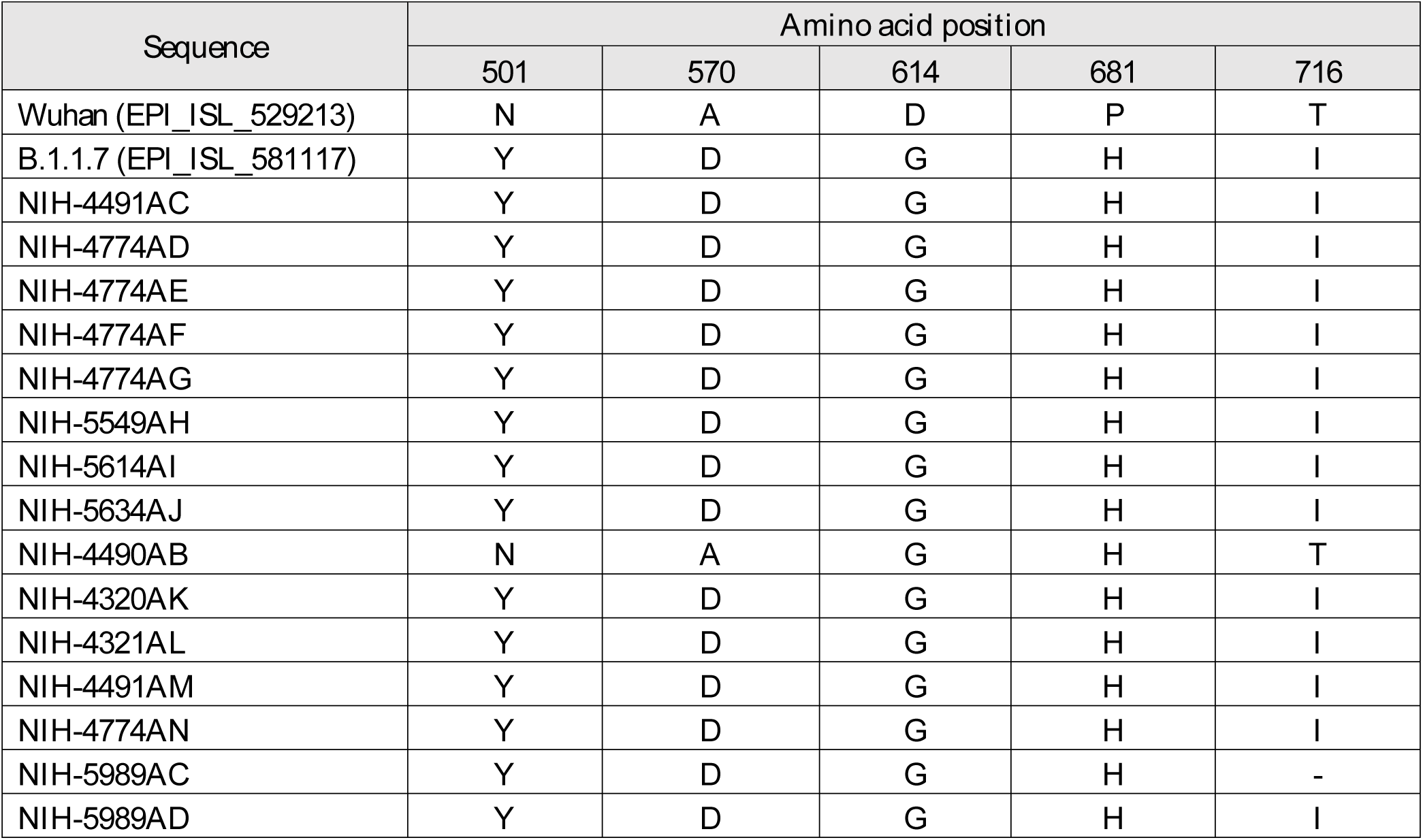
Characteristics mutations of B 1.1.7 variant of concern in the partial spike gene sequence.

In the current study, we found a sharp rise in SGTF samples in February 2021, an indicator of the rapid spread of B1.1.7 variant in the indigenous population. The use of SGTF through ThermoFisher TaqPath™ kit for the detection of B.1.1.7 has been successfully used in the UK where more than 97% of SGTF samples were confirmed to be the B.1.1.7 [15]. Although the SGTF provides a practical option for the screening of the B.1.1.7 variant particularly in resource poor setting where genomic surveillance is not possible, it is not definitive for B.1.1.7 [16, 17]. Therefore, we used partial sequencing of S gene on a subset of SGTF samples as a confirmatory method for B.1.1.7. The sequencing results confirmed 93.3 % (n=14/15) cases as B.1.1.7 all having the marker S gene mutations (N501Y, A570D, P681H, and T716I).

The small size representative of a defined geographic area is a major limitation of the study. However, our findings highlight the possible spread of the B.1.1.7 in indigenous population and warrants large scale genomic surveillance. Moreover, it will guide health authorities to take appropriate measures to prevent further spread of the variant in the country.

## Data Availability

All the data has been made available
Genetic sequences have been submitted in GISAID

https://www.gisaid.org/

## Data Availability

All the data has been made available
Genetic sequences have been submitted in GISAID

https://www.gisaid.org/

We declare no competing interests.

## Notes

### Competing Interest Statement

The authors have declared no competing interest.

### Funding Statement

No funding was received for any part of this work

### Author Declarations

Institutional Review Board of National Institute of Health, Pakistan

